# Quantitative assessment on the severity degree of Alzheimer dementia by algebraic analysis on cortical thickness profiles of human brains

**DOI:** 10.1101/2020.10.13.20212324

**Authors:** Sangyeol Kim, Seongjun Park, Iksoo Chang, the Alzheimer’s Disease Neuroimaging Initiative

**Author notes:** Contributed equally to this work (Sangyeol Kim, Seongjun Park). Correspondence: Sangyeol Kim and Iksoo Chang. Data used in preparation of this article were obtained from the Alzheimer’s Disease Neuroimaging Initiative (ADNI) database (adni.loni.usc.edu). As such, the investigators within the ADNI contributed to the design and implementation of ADNI and/or provided data but did not participate in analysis or writing of this report. A complete listing of ADNI investigators can be found at: http://adni.loni.usc.edu/wp-content/uploads/how_to_apply/ADNI_Acknowledgement_List.pdf.

## Abstract

Alzheimer disease(AD) affects profoundly the quality of human life. Quantifying the severity degree of Alzheimer dementia for an individual person is critical for the early diagnose and prescription for delaying the further dementia progression. However, the quantitative diagnose for human subjects of the mild cognitively impairment(MCI) or AD with the different degree of dementia severity is a difficult task due to both the very broad distribution of dementia severities and the lack of good quantitative determinant to assess it. Here, based on cortical thickness profiles of human brains measured by magnetic resonance experiment, a new algebraic approach is presented for the personalized quantification of the severity degree of AD, ranging from 0 for the basin of cognitively normal state to 1 for AD state. Now one can unfold the broad distribution of dementia severities and corresponding cortex regions of human brains with MCI or AD by the different severity degree.

## Introduction

Alzheimer’s disease (AD) is one of the most well-known neurodegenerative diseases, and it profoundly affects the quality of human life. Various causes of AD are known, and clinical treatments for preventing or delaying the progression of AD are currently practiced. The precise diagnosis of AD requires not only the systematic identification of cohorts that classify the different stages of brain progression toward AD but also the estimation of the severity degree of AD for a given individual (Galvin et al., 2012, Solomon and Soininen, 2015, Raj et al., 2012). The symptoms of AD appear in various forms in the human body, behavior, and cognition, yet the direct anatomical evidences appear in the structural change within the brain cortex (Hojjati et al., 2019, Kim et al., 2020, Qiu et al., 2020, Tetreault et al., 2020). Among these evidences is the degradation of the cortical thickness of the human brain, which is one of the imprints of AD. Such physical change can be monitored through the neuro-image analysis, for example the magnetic resonance image (MRI) analysis of the brain (Hartikainen et al., 2012, Im et al., 2008, Kim et al., 2014, Lebedev et al., 2013, Paternico et al., 2016, Querbes et al., 2009).

The anatomical degradation of the cortical thickness becomes more pronounced as the degree of dementia becomes greater (Im et al., 2008, Querbes et al., 2009). Therefore, there is a rationale for taking advantage of the degree of cortical degradation for determining the dementia cohort and estimating the severity of dementia progression. Given the big data information of cortical thickness for a single subject, however, we are faced with several obstacles to overcome in accomplishing abovementioned tasks. First, we noted that the person-to-person fluctuations in cortical thickness of an individual may overwhelm the degradation in cortical thickness. In clinical cases, we frequently observed that the average cortical thickness of some cognitively normal people is thinner than that of people with AD, which appears to contrast conventional view. The second obstacle is the ambiguity in what we should do if two different cohorts have a difference in the cortical thickness in brain regions that have little to do with AD. In principle, we should construct some determinants for distinguishing a cognitively normal person from a person with AD based on cortical thickness, but in practice, we are confronted with the differences in cortical thickness in regions of the cortex that are unrelated to the pathogenesis of AD. We noted that the abundant existence of such unrelated regions is an intrinsic source that increases the uncertainty of the dementia determinants and hinders the appropriate construction of good classifier and predictor for AD.

In this study, instead of dealing with all 327,684 vertices point on the whole cortex of a human brain, we demonstrated that the consideration of a few hundred essential vertices were enough for distinguishing CN, MCI, AD cohorts each other. With cortical thickness data at these essential vertices, we defined the statistical score matrix and the covariance correlation matrix between human subjects as a set of classifier and predictor for AD. Unlike the conventional view that the degradation of the cortical thickness of human brain was responsible for AD, the singular valued decomposition analysis of the statistical score matrix developed in this study implicated that the simultaneous consideration of both thinner and thicker cortical regions together compared to those of CN are important and necessary for the precise diagnose of the severity of AD.

## Materials and methods

### Preparation of cortical thickness data from MRI of 1522 human brain images from ADNI

Data used in the preparation of this article were obtained from the Alzheimer’s Disease Neuroimaging Initiative (ADNI) database (adni.loni.usc.edu). The ADNI was launched in 2003 as a public-private partnership, led by principal investigator Michael W. Weiner, MD. The primary goal of the ADNI has been to test whether serial MRI, PET, other biological markers, and clinical and neuropsychological assessment can be combined to measure the progression of MCI and early Alzheimer’s disease. For up-to-date information, see www.adni-info.org.

We selected 274 individuals (human subjects) who were identified as CN, 265 individuals with MCI, 125 individuals with AD from the ADNI-2 study of ADNI, and 97 with MCI from the ADNI-GO study of ADNI. A human brain image-data set of 1522 MR images from a total of 761 subjects was constructed, for each of which both 1.2-mm sagittal Magnetization Prepared Rapid Gradient Echo (MPRAGE) and MPRAGE_SENSE2 images were taken separately.

### Partition 1516 MR images of human brains into four groups and determine the essential region-of-interest vertices for each group

We performed the FreeSurfer analysis to obtain the cortical thickness data at 327,684 vertices on the cortex of a human brain (Dale et al., 1999, Fischl et al., 1999). The cortical thickness at each vertex ranges from 0 mm to 5 mm. After eliminating those vertices at which cortical thickness information was missing for any one of the 1522 MR images of human brains in the ADNI data set, 276,825 common vertices whose cortical thickness values are available for all 1522 MR images were selected for our study. The average cortical thickness over 276,825 vertices for each brain images was evaluated, and we divided 1516 values of average thickness into four groups (A-D) of different windows of average thickness except 6 values of that run out-of-bounds. Demographic characteristics of the average cortical thickness of the four groups are listed in Table 1.

**Table 1.**
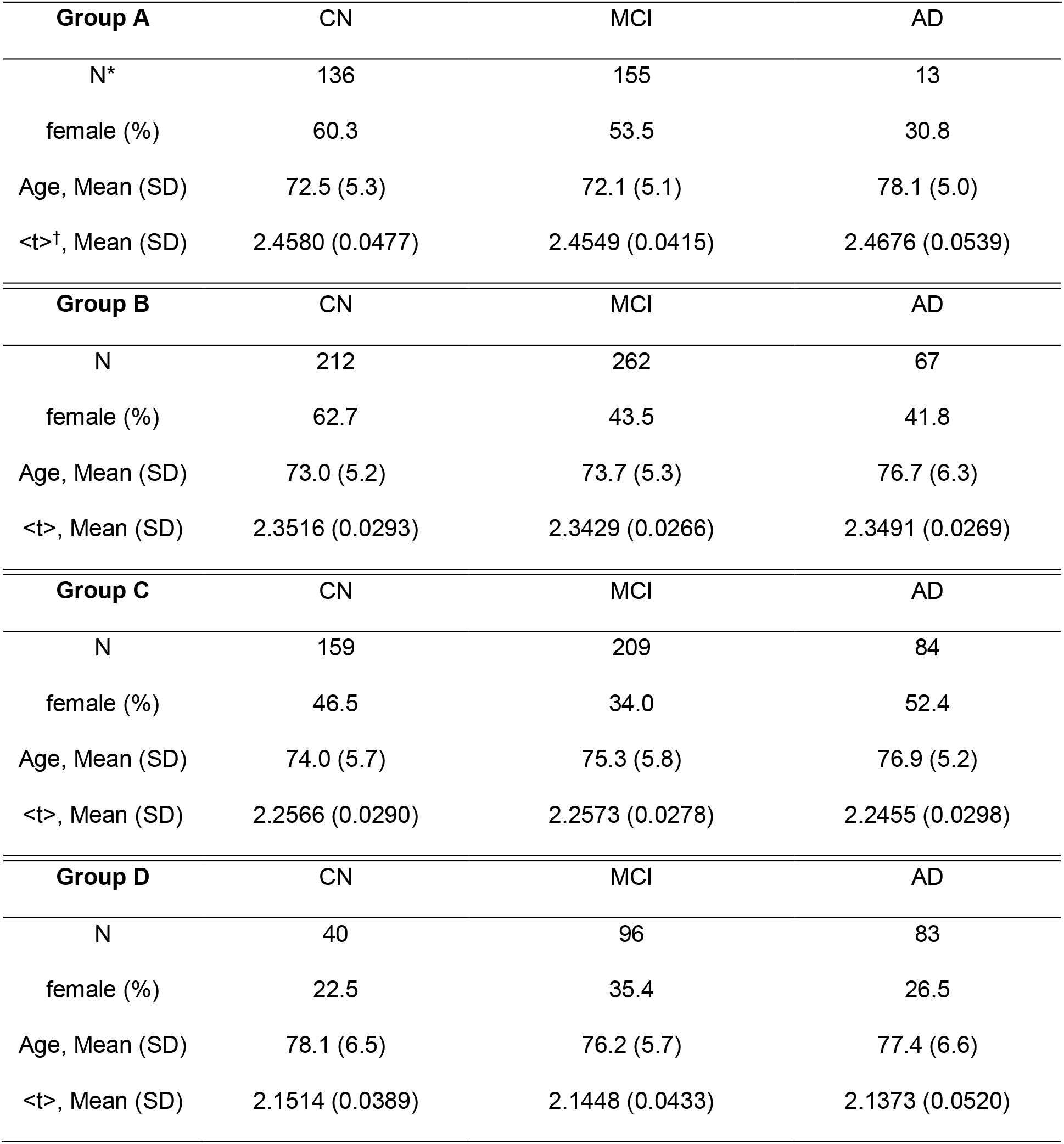
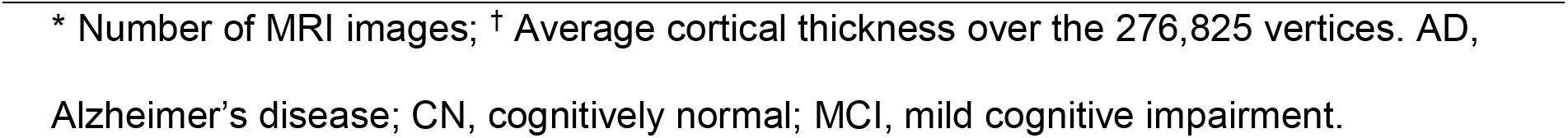
Demographic Characteristics of the groups.

| **Group A** | CN | MCI | AD |
| --- | --- | --- | --- |
| N* | 136 | 155 | 13 |
| female (%) | 60.3 | 53.5 | 30.8 |
| Age, Mean (SD) | 72.5 (5.3) | 72.1 (5.1) | 78.1 (5.0) |
| <t><sup>†</sup>, Mean (SD) | 2.4580 (0.0477) | 2.4549 (0.0415) | 2.4676 (0.0539) |
| **Group B** | CN | MCI | AD |
| N | 212 | 262 | 67 |
| female (%) | 62.7 | 43.5 | 41.8 |
| Age, Mean (SD) | 73.0 (5.2) | 73.7 (5.3) | 76.7 (6.3) |
| <t>, Mean (SD) | 2.3516 (0.0293) | 2.3429 (0.0266) | 2.3491 (0.0269) |
| **Group C** | CN | MCI | AD |
| N | 159 | 209 | 84 |
| female (%) | 46.5 | 34.0 | 52.4 |
| Age, Mean (SD) | 74.0 (5.7) | 75.3 (5.8) | 76.9 (5.2) |
| <t>, Mean (SD) | 2.2566 (0.0290) | 2.2573 (0.0278) | 2.2455 (0.0298) |
| **Group D** | CN | MCI | AD |
| N | 40 | 96 | 83 |
| female (%) | 22.5 | 35.4 | 26.5 |
| Age, Mean (SD) | 78.1 (6.5) | 76.2 (5.7) | 77.4 (6.6) |
| <t>, Mean (SD) | 2.1514 (0.0389) | 2.1448 (0.0433) | 2.1373 (0.0520) |

\* Number of MRI images; † Average cortical thickness over the 276,825 vertices. AD, Alzheimer's disease; CN, cognitively normal; MCI, mild cognitive impairment.

In order to assign subjects from each CN, MCI, and AD cohort into one of the four groups (A-D) of average cortical thickness, we employed the *Z* score criteria in selecting the region-of-interest (ROI) vertices and the essential ROI vertices on the cortex at which the distribution of cortical thickness of the CN cohort is distinguished from that of the AD cohort within each group of average cortical thickness. A similar procedure is repeated for distinguishing the CN cohort from the MCI cohort and also the MCI cohort from the AD cohort:

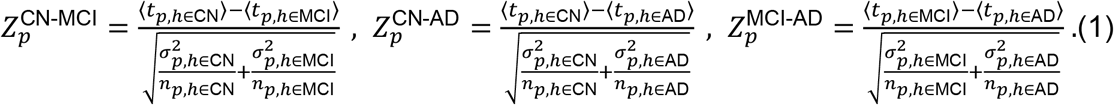

Here, ⟨*t*_*p,h*∈*k*_⟩ is the average cortical thickness at a vertex point *p* averaged over the subject *h* who belongs to the *k* (one of CN, MCI, AD) cohort, and σ_*p,h*∈*k*_ is its standard deviation, and *n*_*p,h*∈*k*_ is the number of MR images belonging to the *k* cohort. The positive (negative) value of 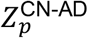, for example, indicates that the distribution curve of the average cortical thickness of the CN cohort is right (or left)-shifted compared to that of AD cohort. And the bigger the absolute value of the *Z* score, the better distinguished the distribution curves of average cortical thickness of the cohorts. In this study, we identified ROI vertices satisfying the absolute value of the *Z* score larger than 1.5, and essential ROI vertices satisfying much higher cut-off Z scores (Table S1).

### Construction of a statistical score matrix for classifying subjects into one of CN, MCI, AD cohorts

Within each group of average cortical thickness, we constructed the statistical score matrix for determining a subject’s cohort as either CN, MCI, or AD (Yu et al., 2011). First of all, *t*_*p,h*∈*k*_ was transformed into the probability distribution matrix 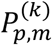, which is a probability that the cortical thickness *t*_*p,h*∈*k*_ at a vertex point *p* of the subject in *k* cohort is between (*m* − 1)*Δ* and *mΔ*:

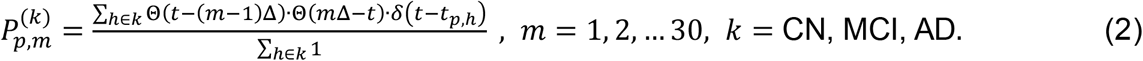

Here, Δ = 0.2 mm, and cortical thickness index *m* runs from 1 to 30; this covers the cortical thickness from 0 mm to 6 mm. δ(x) is a Dirac delta function, and Θ(x) is a step function whereΘ(x < 0) = 0; Θ(x ≥ 0) = 1. Then, we defined the statistical score matrix 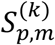 from 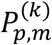 in the following way:

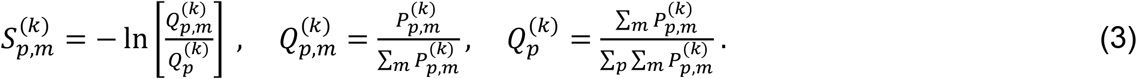

Since 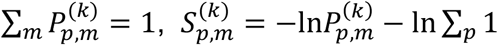 and the second term are constants. The value of the statistical score matrix 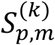 varies depending on the cohort *k*; the smaller 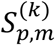 is, the larger 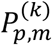 is.

With this statistical score matrix 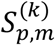, we employed a strategy for determining to which one of *k* cohorts a given subject would belong. First, we evaluated the averaged cortical thickness of a given subject over 276,825 vertices. Second, we assigned this subject to one of four groups (A-D) of average cortical thickness. Third, based on the preselected essential ROI vertices *p* for the assigned group, we determined the cortical thickness index m’(p) at which the cortical thickness at an essential ROI vertex *p* is between (*m* − 1)*Δ* and *mΔ*. Then, for each *k* cohort, the total score 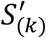 was calculated by summing up 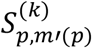 over the preselected essential ROI vertices *p* for the assigned group of the average cortical thickness, 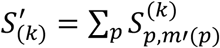. Lastly, to which *k* cohort a given subject would belong was decided by a cohort which gives the minimum score out of S’_(CN)_, S’_(MCI)_, S’_(AD)_.

We, however, noted that the accuracy of both 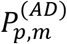 and 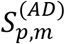 may become unsatisfactory if the number of people in the AD cohort was less than that of the CN cohort and the MCI cohort (Table 1). In order to overcome the unsatisfactory nature of both 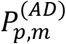 and 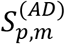, we employed the method of Kernel Density Estimation (KDE); namely, a Dirac delta function δ(*t* − *t*_*p,h*_) in the definition of the probability distribution matrix 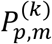, is replaced by a kernel function *f*(t − *t*):

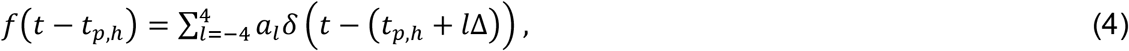

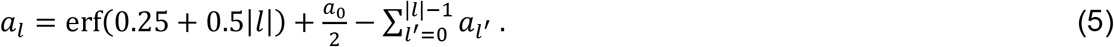

Here, the relative ratio among coefficients *a*_*l*_ is *a*_0_: *a*_±1_: *a*_±2_: *a*_±3_: *a*_±4_ = 56: 43: 21: 7: 1. The kernel function *f*(t − *t*_*p,h*_) satisfies 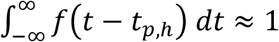 and the standard deviation σ_*f*_ ≈ 1.435. Upon subjecting KDE, 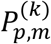, becomes

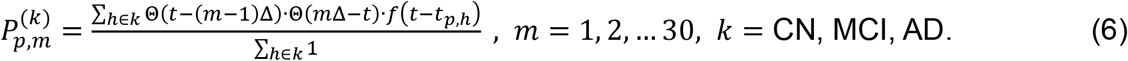

In this study, we constructed the statistical score matrix on which KDE was employed and used it for determining to which *k* cohort a given subject would belong.

### Construction of a covariance correlation matrix and a predictor for the severity degree of AD

Within each group of the average cortical thickness, the severity degree of AD for a given subject is estimated by the following strategy. First of all, we transformed the cortical thickness matrix *t*_*p,h*_ at essential ROI vertices *p* for a subject *h* into the normalized matrix *t*′_*p,h*_ such that

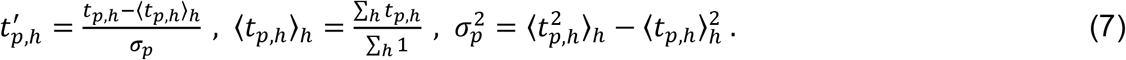

Here, the product of *t*′_*p,j*_ by its transpose *t*′_*p,i*_^*T*^ results in the square matrix *t*′′_*i,j*_ = *t*′_*p,i*_^*T*^·*t*′_*p,j*_, and then its normalized matrix (called by a covariance correlation matrix) *C*_*ij*_ is defined by *C*_*ij*_ = *t*′′_*ij*_/ max{*t*′′_*ij*_}, where max{*t*′′_*ij*_} is the maximum value of elements in the square matrix *t*′′_*ij*_. The larger the value of *C*_*ij*_, the higher the covariance correlation between a subject *i* and a subject *j* in their profile of the cortical thickness at essential ROI vertices. Based on this covariance correlation matrix, we defined the severity degree AD for a given subject *i by*

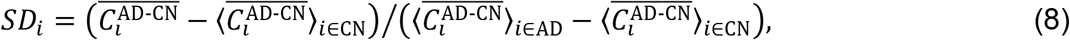

Where 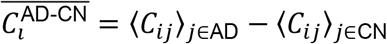. The severity degree of AD ranged from 0 for the basin of CN state to 1 for the basin of AD state. Rank-ordering this degree in ascending order illustrates that a subject *i* with the larger (or smaller) value of the severity degree is more prone to AD (CN) state.

## Results

### Identification of essential ROI vertices at which the distributions of cortical thickness of CN, MCI, AD subjects are distinguishable

Although the averaged cortical thickness of subjects with AD is generally known to be thinner than that of CN or MCI subjects, the distribution curves of averaged cortical thickness for the cohorts are not well distinguishable except near both ends of the distribution curves as demonstrated in Figure 1A. This illustrates that a subject can be CN even though the averaged cortical thickness is thinner than that of a subject with AD, and vice versa. Also, we found that many subjects identified as CN, MCI, or AD have a similar averaged cortical thickness. This is due to the fact that the average cortical thickness for a subject was calculated over all 276,825 vertices, and the cortical thicknesses at most vertices are similar for all subjects, which prohibits us from successfully clustering 1552 human brain images into the image of CN, MCI, AD cohorts. Therefore, instead of resorting on the cortical thickness of all 276,825 vertices, we need to select the ROI vertices at which the cortical thickness values are distinguishable from each other among CN, MCI, and AD. For a fair selection of such ROI vertices, we divided the range of the averaged cortical thickness of subjects into four (A-D) different regions (for the detailed method, see in the section 2.2). Figure 1B illustrates how we identified ROI and essential ROI vertices. The x-axis 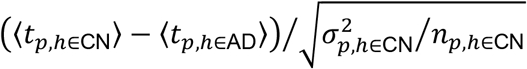 represents the degree of separation between the distribution curves of cortical thickness for CN subjects and AD subjects at a vertex *p* normalized by the dispersion of the cortical thickness of CN subjects, which is quantified by the value of its *Z* score. The y-axis 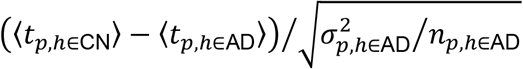 represents values normalized by the dispersion of the cortical thickness of individuals with AD. Therefore, the x-values (y-values) at a vertex point *p* represent the degree by which the distribution of cortical thickness at this point *p* of CN (AD) subjects is distinguished from averaged cortical thickness of subjects with AD (CN). It means the larger the value of 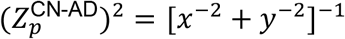 is, the two distribution curves are more distinguished each other (for the illustration, see Figure S1). The ROI cut-off line is defined by points satisfying 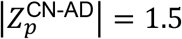, and the distribution of cortical thickness of CN subjects and individuals with AD is clearly distinguished at those points satisfying 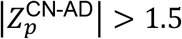 (outside of the ROI cut-off line).

**Figure 1.**
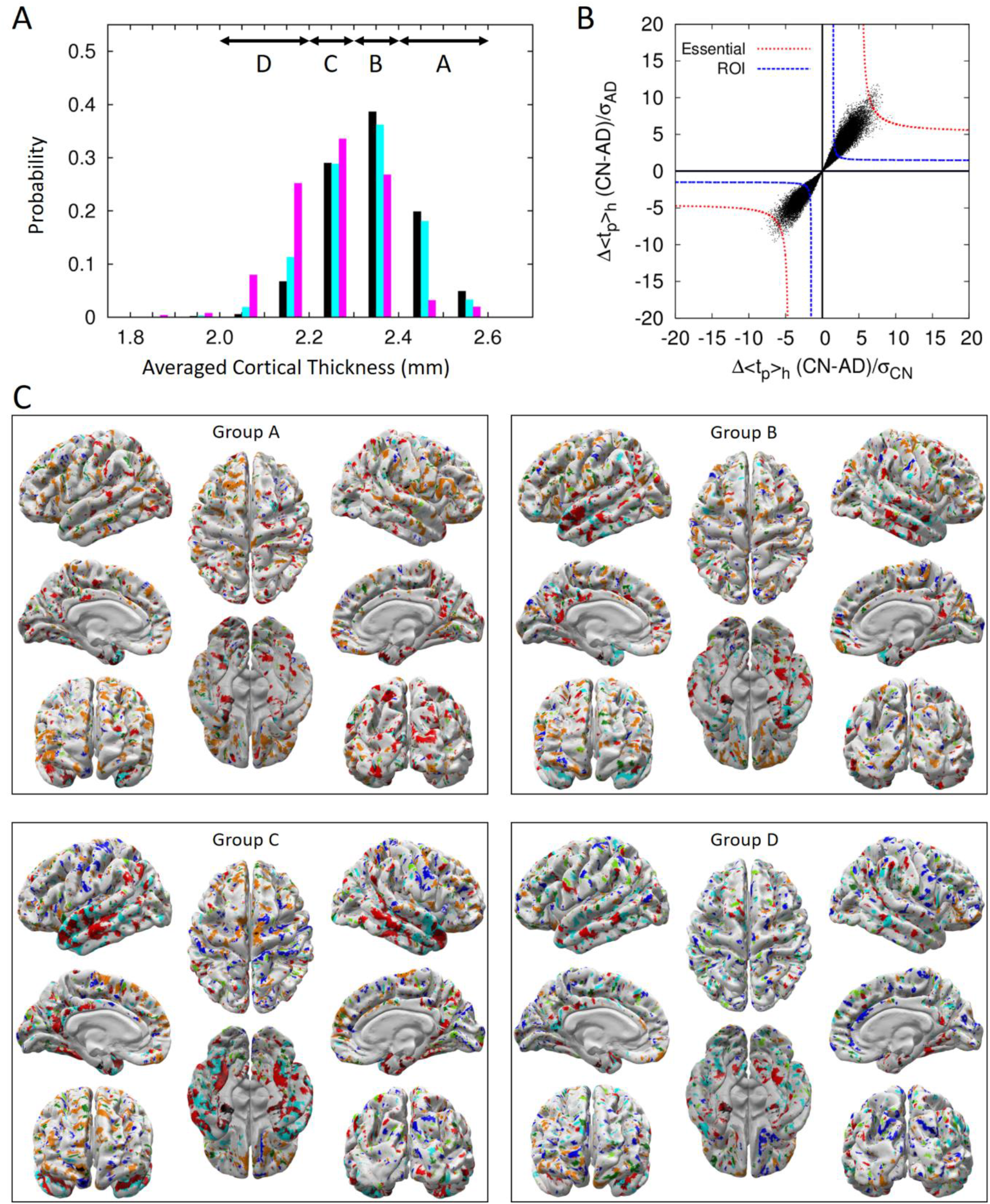
The classification of 1516 brain images into four groups by the average cortical thickness of subjects from the ADNI. (A) The distribution of average cortical thickness of subjects in the CN (black), MCI (cyan), and AD (magenta) cohorts. Above arrows point to the range of average cortical thickness. Subject number, sex, and age for each group are listed in Table 1. (B) For the cortical thickness group D, the degree of separation of the distribution curve of average cortical thickness between CN subjects and AD subjects is presented in the form of black points. The closer to the origin point (0, 0) the degree of separation of two distribution curves of average cortical thickness is, the less distinguishable they are (Figure S1). Black points residing outside of the blue-dashed line (*Z* = ±1.5) are ROI vertices, and black points residing outside of red-dashed line (*Z* values are listed in Table S1) are essential ROI vertices. (C) For each group of average cortical thickness, ROI vertices at which the thickness of the cortex for CN subjects is thicker (thinner) than that of the other subjects with MCI or AD is represented by cyan (blue) color. As a similar procedure, ROI vertices for MCI subjects is thicker (thinner) than the other cohorts is represented by green (dark green) color. Also, ROI vertices for AD subjects is thicker (thinner) than the other cohorts is represented by orange (red) color. Especially, the ROI vertices at which the cortical thickness decreases in the descending order of CN-MCI-AD is represented by dark red. And the essential ROI vertices are represented by a black color. The ROI vertices commonly found from more than three groups of average cortical thickness are presented in Figure S2.

Figure 1C shows ROI vertices for each of the cortical thickness groups (A-D) by colored points on the white cortex, at which the thickness of the cortex is either thicker or thinner particularly for one cohort compared with that of two other cohorts. These ROI vertices are widely distributed on the cortex, and their locations are not fixed but vary depending on the groups A to D. We uncovered, however, that the medial temporal lobe, known to be very important for memory formation, is always indicated by a red or dark red color irrespective of the groups A to D (Figure S2). This implies that the cortical thickness values of the medial temporal lobe for subjects with AD are characteristically thinner than those of CN subjects or individuals with MCI (red color), and this decrease occurs in the following descending order: CN-MCI-AD (dark red color). The medial temporal lobe is the region where the cortical thickness gradually decreases as dementia progresses and therefore is the critical region necessary for determining the dementia cohort and the severity degree of AD. We also noted that the cortical thickness at the orange-colored region for subjects with AD is thicker than that for CN subjects or those with MCI. This has nothing to do with the damage in the cortex but contributes to the increase in the accuracy for predicting the dementia cohort since it could provide better distinguishability of subjects with AD from CN subjects and those with MCI.

### Character of statistical score matrix and classification of subject’s cohort

The section 2.3 described the detailed procedure of constructing the statistical score matrix for determining a subject’s cohort within each group of the average cortical thickness (Figure 2A). In order to judge how well the statistical score matrix distinguishes CN, MCI, and AD cohorts from each other before we predict the cohort of a new subject, we performed the singular value decomposition (SVD) analysis on the combined statistical score matrix S^(All)^ which consists of matrix elements of S^(CN)^, S^(MCI)^, and S^(AD)^. We used the SVD character of a matrix that a given matrix can be reconstructed as a linear combination of the products of two singular vectors by one singular matrix. Since the reconstructed matrix from the few highest modes of singular values contains the predominant character of an original given matrix, one expects that the differences among the cohorts should appear in singular vectors of different cohorts. Otherwise, the statistical score matrix S^(All)^ is not reliable nor does it contain the characteristic ingredient of different cohorts. Figure 2B, 2C and S3 show the highest six singular vectors corresponding to the six largest singular values from SVD analysis of the statistical score matrices for each group of the average cortical thickness A to D. Here, it demonstrates that elements in the singular vectors *v*_1_ to *v*_3_ for CN, MCI, and AD follow qualitatively a similar trend, meaning that these compose the fundamental default modes, whereas those in *v*_4_ to *v*_5_ follow a different trend and are distinguished each other.

**Figure 2.**
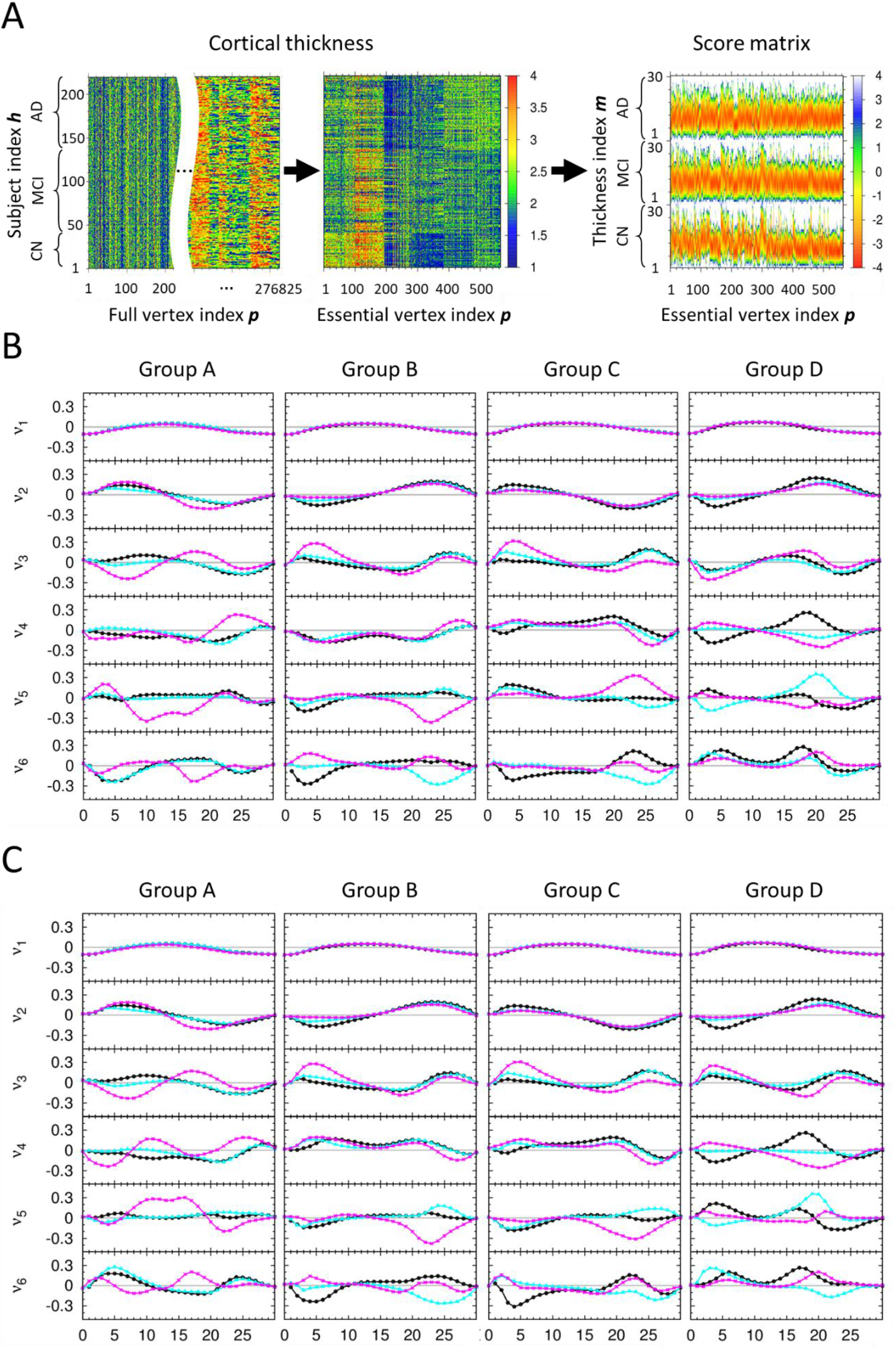
The character of score matrix for each group of average cortical thickness. (A) For a given group of average cortical thickness, three kinds of heat maps illustrate the process starting from the cortical thickness matrix at all 276,825 vertices to that at only 564 essential ROI vertices, and then construction of the score matrix. The dimension in the x-axis of the cortical thickness matrix at all 276,825 is too large to draw, we placed the blank in the middle to abbreviate the large dimension of the x-axis. (B) The results of singular value decomposition analysis on score matrices, which are composed of 547 CN, 722 MCI, 247 AD human brain images and used for self-recognition test. For each group of average cortical thickness, six singular vectors corresponding to the six largest singular values are presented. Here, x-axis is *m* value defined in the section 2.3, and y-axis is an arbitrary unit for the singular vectors. For each graph, the singular vector components for CN, MCI, and AD subjects are plotted by black, cyan, magenta colors, respectively. (C) The results of singular value decomposition analysis on score matrices, which are composed of 363 CN, 480 MCI, 163 AD human brain images as a training set and used for the first iteration of the stratified 3-fold cross validation test. The other results of that used for the second and third iterations of the stratified 3-fold cross validation test are presented in Figure S3.

Out of 547 CN, 722 MCI, and 247 AD human brain images predetermined clinically and provided from the ADNI data set, we performed the self-recognition test and the stratified 3-fold cross validation test using the 1516 human brain images for each group on the average cortical thickness A to D (Table 2). For the first (second; third) iteration of stratified 3-fold cross validation test, 1006 (1011; 1015) human brain images were used as the training set for learning the statistical score matrix and 510 (505; 501) human brain images were used as an independent validation set. The new method presented in this study recognized and predicted the subjects with AD in the cohort with more than 91% (self-recognition test) and 82% (stratified 3-fold cross validation test) accuracy, respectively.

**Table 2.** Result for the tests of the cohorts for each group.

| Self-recognition test |  |  |  |  |  |  |  |  |  |  |  |  |  |  |
| --- | --- | --- | --- | --- | --- | --- | --- | --- | --- | --- | --- | --- | --- | --- |
| Exp. | Score | Group A |  |  | Group B |  |  | Group C |  |  | Group D |  |  | Correct (%) |
|  |  | CN | MCI | AD | CN | MCI | AD | CN | MCI | AD | CN | MCI | AD |  |
| CN |  | 128 | 8 | - | 197 | 12 | 3 | 142 | 11 | 6 | 40 | - | - | 507 (92) |
| MCI |  | 16 | 139 | - | 74 | 155 | 33 | 59 | 120 | 30 | 5 | 88 | 3 | 502 (69) |
| AD |  | - | - | 13 | 6 | 1 | 60 | 7 | 2 | 75 | 1 | 3 | 79 | 227 (91) |

| Stratified 3-fold cross validation test |  |  |  |  |  |  |  |  |  |  |  |  |  |  |
| --- | --- | --- | --- | --- | --- | --- | --- | --- | --- | --- | --- | --- | --- | --- |
| Exp. | Score | Group A |  |  | Group B |  |  | Group C |  |  | Group D |  |  | Correct (%) |
|  |  | CN | MCI | AD | CN | MCI | AD | CN | MCI | AD | CN | MCI | AD |  |
| CN |  | 122 | 14 | - | 177 | 28 | 7 | 133 | 20 | 6 | 31 | 6 | 3 | 463 (84) |
| MCI |  | 33 | 122 | - | 97 | 130 | 35 | 68 | 103 | 38 | 9 | 75 | 12 | 430 (59) |
| AD |  | 1 | - | 12 | 5 | 9 | 53 | 7 | 11 | 66 | 2 | 8 | 73 | 204 (82) |

“Exp.” column, outside of the parenthesis, represents the number of MR images base on the clinical test, and “Score” row, inside of the parenthesis, represents the number of MR images base on our test using score matrix. AD, Alzheimer’s disease; CN, cognitively normal; MCI, mild cognitive impairment.

### Estimating the severity of AD of subjects by covariance correlation matrix

Developing a quantitative measure to tell how serious the degree of dementia progression for a given subject is very important for diagnosing and clinically treating patients with MCI and AD with the different degree of Alzheimer dementia. In this study, we already identified essential ROI vertices and constructed the statistical score matrices as a classifier ensuring the correct prediction of subjects with AD at more than 80% accuracy that they belong to the AD cohort. Thus, we extracted the cortical thickness profile (or vector) at essential ROI vertices for all brain images, and constructed the covariance correlation matrix between them. Then we compared the profile for a given subject’s image with that of patients with AD, to estimate the severity degree of AD for a given subject towards patients with AD (for the detailed method, see the section 2.4 and Figure 3A). The personalized quantitative severity degree of dementia (see the equation (8)) is plotted at the right-bottom graph of Fig. 3A for each subject of CN, MCI, AD cohort of the group D in the ascending order. The values of severity degree of dementia were distributed around the averaged value of 0 (ranging from about −0.5 to +0.5) for subjects with CN, 0.5 (ranging from about −0.2 to +1.2) for subjects with MCI, and 1.0 (ranging from about +0.2 to +1.5) for subjects with AD, respectively. The distribution of the severity degree for subjects with MCI was laid across both ranges of those for CN and AD, which points out that this is the intrinsic source of the low success ratio in determining the dementia cohort of subjects with MCI. One could sort out quantitatively the broad spectrum of the severity of dementia for MCI subjects in that whether they are prone to CN or how much they are progressed toward AD. Given a new person for diagnosing the dementia state, one of the cohort CN, MCI, AD was assigned by the equation (3) and the personalized quantitative severity degree of dementia was estimated by the equation (8). Then, with these two-qualitative and quantitative-determinants, one may infer that a new person with the estimated severity degree below 0.0 is most likely to be CN, with that between 0.0 and 0.5 might be CN or MCI prone to CN, with that between 0.5 and 1.0 might be MCI prone to AD or AD, and with that above 1.0 is most likely to be AD state.

**Figure 3.**
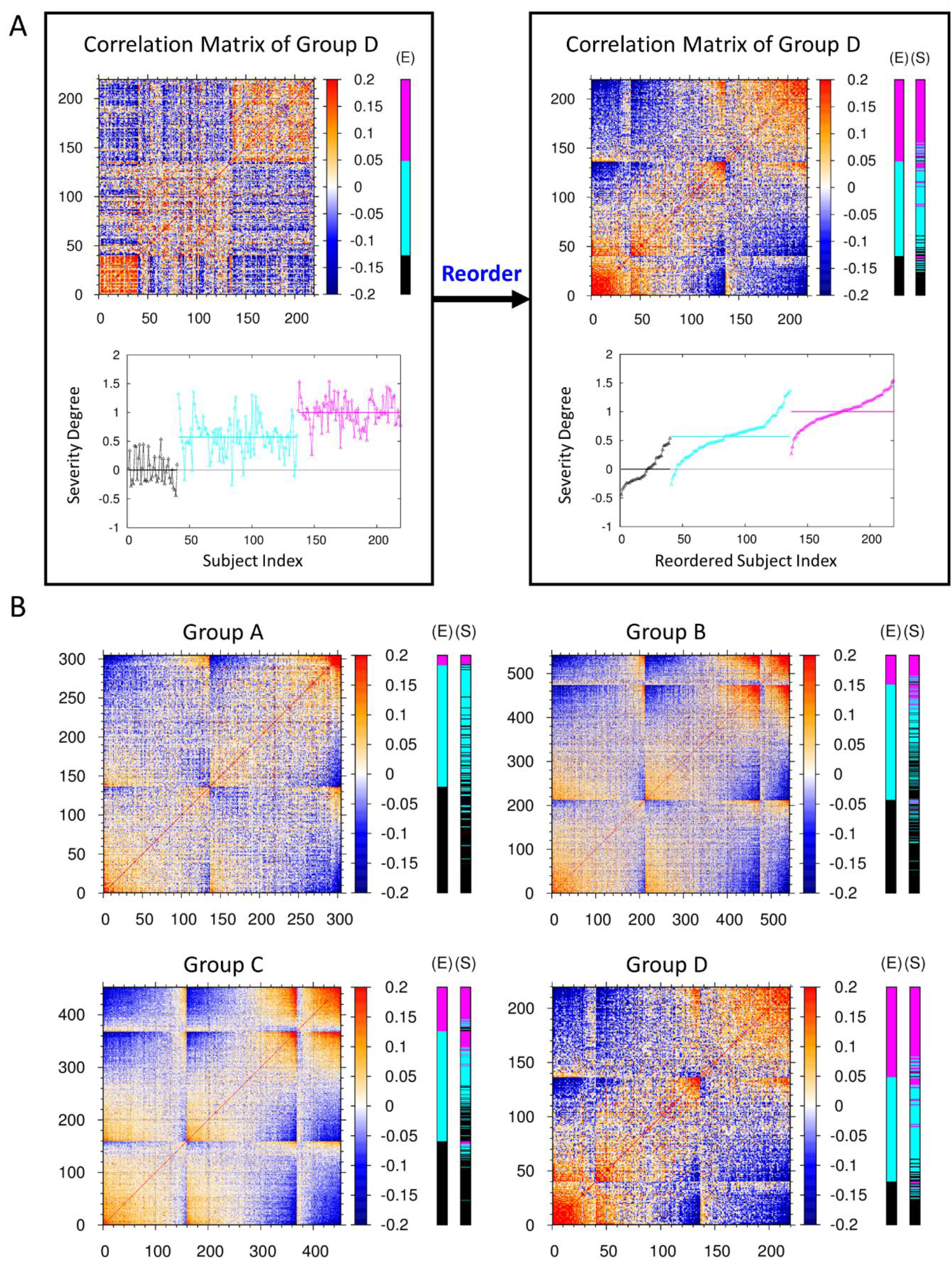
Covariance correlation matrix and severity degree of AD. (A) The left-top heat map is the covariance correlation matrix for group D of average cortical thickness. The x- and y-axes represent the indices 1 to 40 for CN subjects, 41 to 136 for MCI subjects, and 137 to 219 for AD subjects. Here, red (blue) color represents the high (low) correlation between two subjects at the essential ROI vertices. The extra E-cohort color bar at the right of the heat map represents the clinically determined cohort of CN subjects and subjects with MCI and AD denoted by black, cyan, and magenta colors, respectively. The left-bottom graph illustrates the personalized severity degree of dementia for each subjects of the group D in terms of a quantitative value, ranging from 0 for the basin of CN state to 1 for the basin of AD state (for the detailed method, see the section 2.4). The average values of this severity degree in each cohort are denoted by horizontal lines, respectively. The left panel is reordered into the right panel according to the ascending value of the severity degree in each cohort. For those subjects with MCI, the distribution of severity degree of dementia is very broad. One can sort out the broad spectrum of the severity of dementia for MCI subjects in that whether they are prone to CN or how much they are progressed toward AD. (B) The reordered covariance correlation matrices for A, B, C, and D groups of average cortical thickness together with the determination of CN (black), MCI (cyan), and AD (magenta) cohorts by clinical (E-cohort color bar) exam and by the stratified 3-fold cross validation test of this study (S-cohort color bar). The original covariance correlation matrices for the four groups of average cortical thickness are provided in Figure S4.

We constructed the covariance correlation matrices for all groups A, B, C, D of the average cortical thickness and observed the common pattern in the matrices that subjects with AD (CN), possessing a strong correlation among themselves are clustered at the top-right (bottom-left) corner, represented by the cluster of red colors (Figure S4). Also, we presented the reordered covariance correlation matrices by the severity degree of AD and the results to which one of the CN, MCI, AD cohorts each human brain would belong, based on both the clinical test and our stratified 3-fold cross validation test (Figure 3B). After comparing the result from our independent validation test with that of the clinical test, we noted that those subjects which were predicted to belong to the MCI cohort by the clinical test and yet estimated to have the higher (lower) severity degree of AD by our estimation, were predicted to belong to the AD (CN) cohort from the our validation test.

## Discussion

Based on the cortical thickness data of 1516 human brain images from the ADNI data set, we presented a new algebraic approach for both (1) the identification of the cohort (CN, MCI, AD) a given subject would belong to and (2) the quantitative estimation of the severity degree of AD for a given new person (Figure 4). A total of 1516 human brain MR images were partitioned into four groups by the average cortical thickness of each subject. Out of 327,684 vertices on the cortex, a few hundred essential ROI vertices for each group were identified, which were enough to distinguish the cortical thickness distribution of the CN, MCI, and AD cohorts from each other. Statistical score matrices using the cortical thickness on the essential ROI vertices were constructed as a classifier for determining the cohort of a given subject. Out of 547 CN, 722 MCI, and 247 AD subjects predetermined clinically, the success ratio for recognizing their cohort was 92% with CN, 69% with MCI, and 91% with AD subjects. On the other hand, the stratified 3-fold cross-validation test gave the correct prediction rate of 84% with CN, 59% with MCI, and 82% in subjects with AD; this is in agreement with the results of clinical determination. Using the quantitative severity degree of AD for subjects, we could explain the reason why the inevitable uncertainty in the determination of the MCI cohort arouse by the very broad distribution of the severity degree of dementia which MCI subjects possess intrinsically. We suggested that the severity degree of AD presented in this study would be a realistic measure for the quantitative personalized diagnosis of a given subject instead of tri-partitioning the classification of a subject’s cohort only by CN, MCI or AD. It is the continuous degree for a given subject along the scale from 0 for the basin of CN state to 1 for the basin of AD state. One could sort out quantitatively the broad spectrum of the severity degree of dementia for MCI or AD subjects with the different severity degree of dementia in that whether they are prone to CN or how much they are progressed toward AD. This study not only provided a straightforward algebraic approach to analyzing the cortical thicknesses of human brains but also suggested quantitative measures by which one could estimate both the cohort and the severity degree of AD for a given new subject based on the neuro-images from the structural MRI. The MRI data of a larger number of human brains could also be implemented into this study in a systematic and robust manner, which would facilitate the better diagnose of AD with the different degree of dementia.

**Figure 4.**
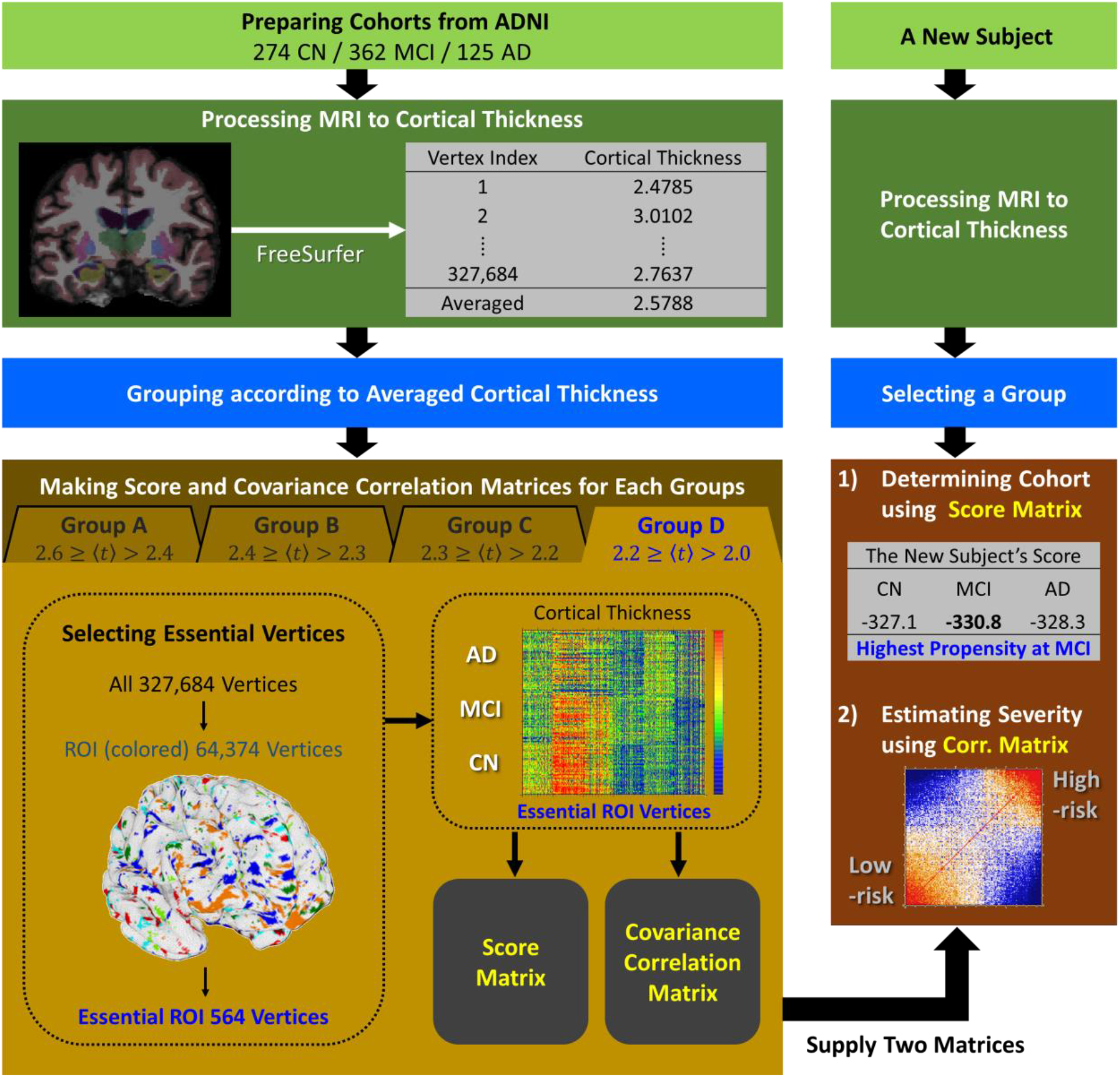
The flow chart for the determination of cohorts and the estimation of the severity degree of AD. The left shows the process of constructing the score matrix and covariance correlation matrix from the cortical thickness big data of subjects derived from the ADNI data set. The right shows the process of determining the cohort and the severity degree of AD for a new given subject.

## Data Availability

Data used in preparation of this article were obtained from the Alzheimers Disease Neuroimaging Initiative (ADNI) database (adni.loni.usc.edu). As such, the investigators within the ADNI contributed to the design and implementation of ADNI and/or provided data but did not participate in analysis or writing of this report. A complete listing of ADNI investigators can be found at: http://adni.loni.usc.edu/wp-content/uploads/how_to_apply/ADNI_Acknowledgement_List.pdf.

## Acknowledgments

This work was supported by the National Research Foundation, Korea [grant number NRF 2017R1E1A1A03070854]. We acknowledged DGIST supercomputing big data center for the allocation of supercomputing resources. We appreciated Mookyung Cheon and Wookyung Yu for the fruitful discussions, and also Keonho Lee and Jangjae Lee of Chosun university for the preparation of MRI image data in the initial stage of this work.

Data collection and sharing for this project was funded by the AD Neuroimaging Initiative (ADNI) (National Institutes of Health Grant U01 AG024904) and DOD ADNI (Department of Defense award number W81XWH-12-2-0012). ADNI is funded by the National Institute on Aging, the National Institute of Biomedical Imaging and Bioengineering, and through generous contributions from the following: AbbVie, Alzheimer’s Association; Alzheimer’s Drug Discovery Foundation; Araclon Biotech; BioClinica, Inc.; Biogen; Bristol-Myers Squibb Company; CereSpir, Inc.; Cogstate; Eisai Inc.; Elan Pharmaceuticals, Inc.; Eli Lilly and Company; EuroImmun; F. Hoffmann-La Roche Ltd and its affiliated company Genentech, Inc.; Fujirebio; GE Healthcare; IXICO Ltd.; Janssen Alzheimer Immunotherapy Research & Development, LLC.; Johnson & Johnson Pharmaceutical Research & Development LLC.; Lumosity; Lundbeck; Merck & Co., Inc.; Meso Scale Diagnostics, LLC.; NeuroRx Research; Neurotrack Technologies; Novartis Pharmaceuticals Corporation; Pfizer Inc.; Piramal Imaging; Servier; Takeda Pharmaceutical Company; and Transition Therapeutics. The Canadian Institutes of Health Research is providing funds to support ADNI clinical sites in Canada. Private sector contributions are facilitated by the Foundation for the National Institutes of Health (www.fnih.org). The grantee organization is the Northern California Institute for Research and Education, and the study is coordinated by the Alzheimer’s Therapeutic Research Institute at the University of Southern California. ADNI data are disseminated by the Laboratory for Neuro Imaging at the University of Southern California.

## Supplementary materials

**Supplementary Table 1.** Upper / lower cut-off *Z* score and number of essential ROI vertices.

| | $Z^{\text{CN-MCI}}$ | $Z^{\text{CN-AD}}$ | $Z^{\text{MCI-AD}}$ | Number of<br>essential ROI vertices |
| --- | --- | --- | --- | --- |
| Group A | 4.4 / -4.1 | 6.3 / -5.0 | 6.4 / -5.1 | 479 |
| Group B | 4.7 / -3.9 | 6.1 / -4.4 | 4.8 / -4.3 | 520 |
| Group C | 4.9 / -4.4 | 8.7 / -4.3 | 5.7 / -4.2 | 494 |
| Group D | 4.2 / -4.2 | 5.5 / -4.6 | 4.7 / -4.2 | 564 |

AD, Alzheimer's disease; CN, cognitively normal; MCI, mild cognitive impairment; ROI, region-of-interest.

**Supplementary Figure 1.**
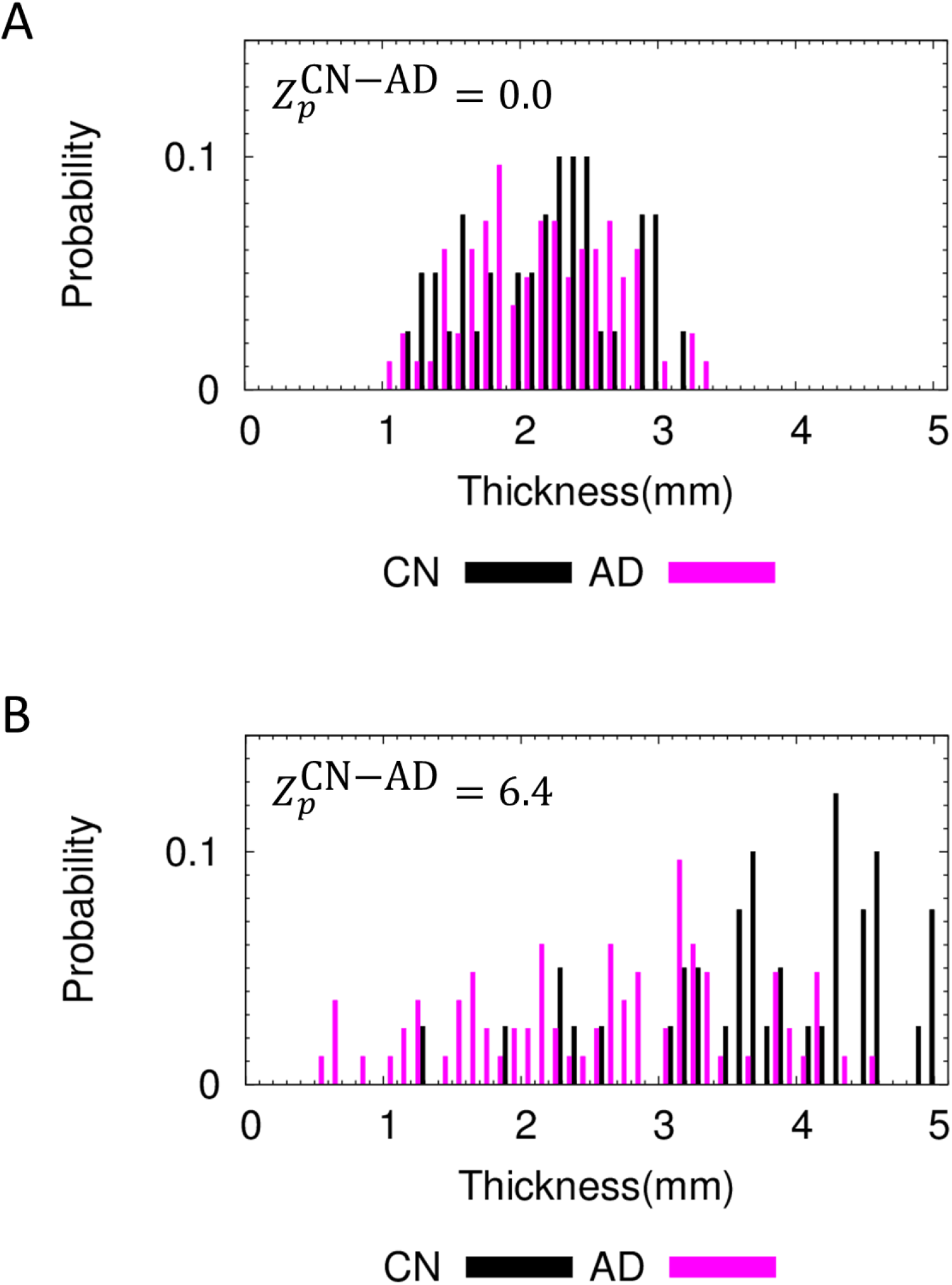
The distribution of cortical thickness of CN and AD subjects in group D of average cortical thickness. (A) At the smallest 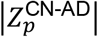 value, the two distribution curves of CN and AD subjects are not distinguished. On the other hand, (B) at the largest 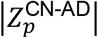 value, the two distribution curves are distinguished.

**Supplementary Figure 2.**
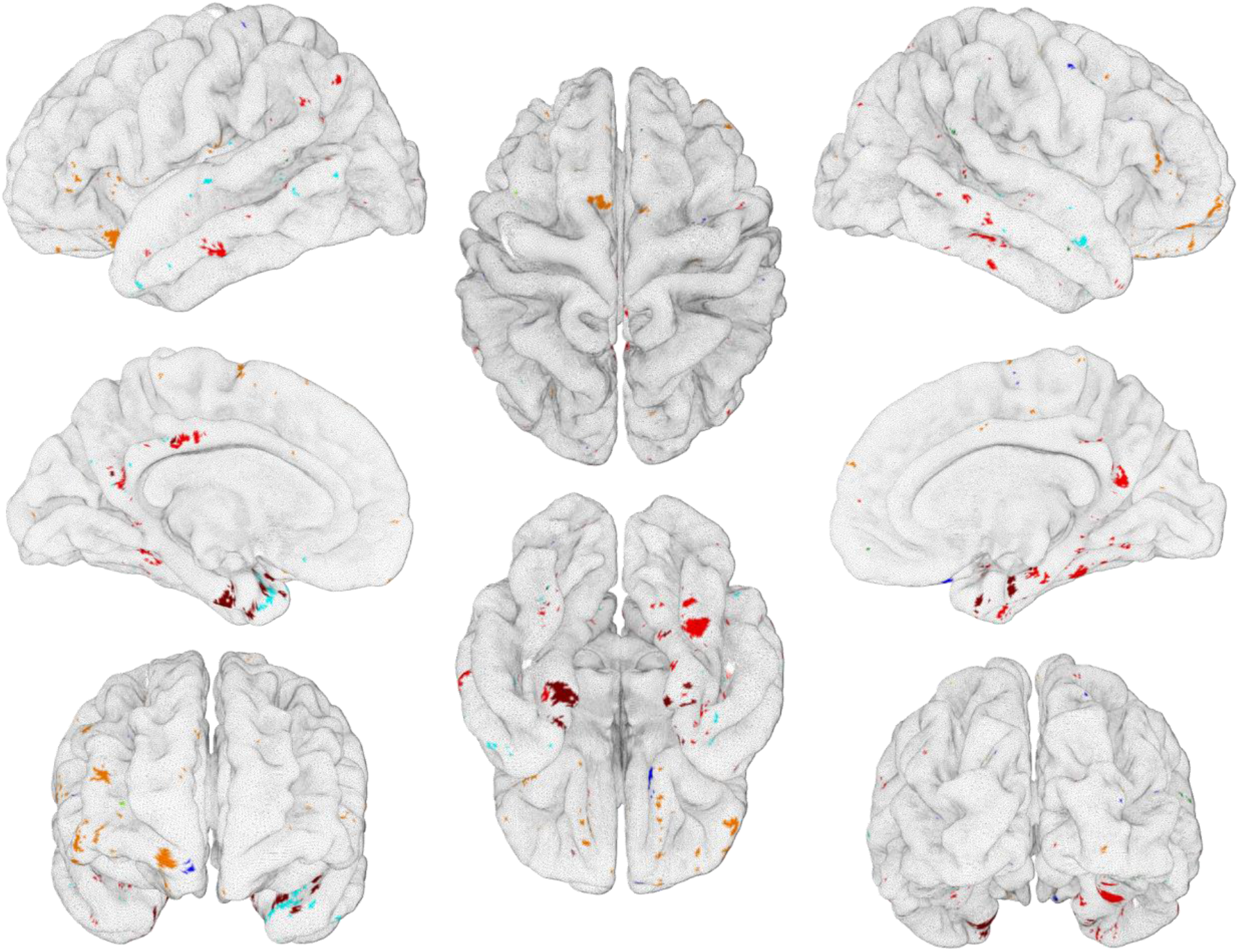
ROI vertices found commonly more than three times from A, B, C, and D groups of the average cortical thickness. ROI vertices at which the cortical thickness of CN subjects are thicker (thinner) are represented by cyan (blue) color. ROI vertices at which the cortical thickness of subjects with MCI are thicker (thinner) are represented by green (dark green) color. ROI vertices at which the cortical thickness of subjects with AD are thicker (thinner) are represented by orange (red) color. The ROI vertices at which the cortical thickness decreases in the descending order of CN-MCI-AD are represented by dark red.

**Supplementary Figure 3.**
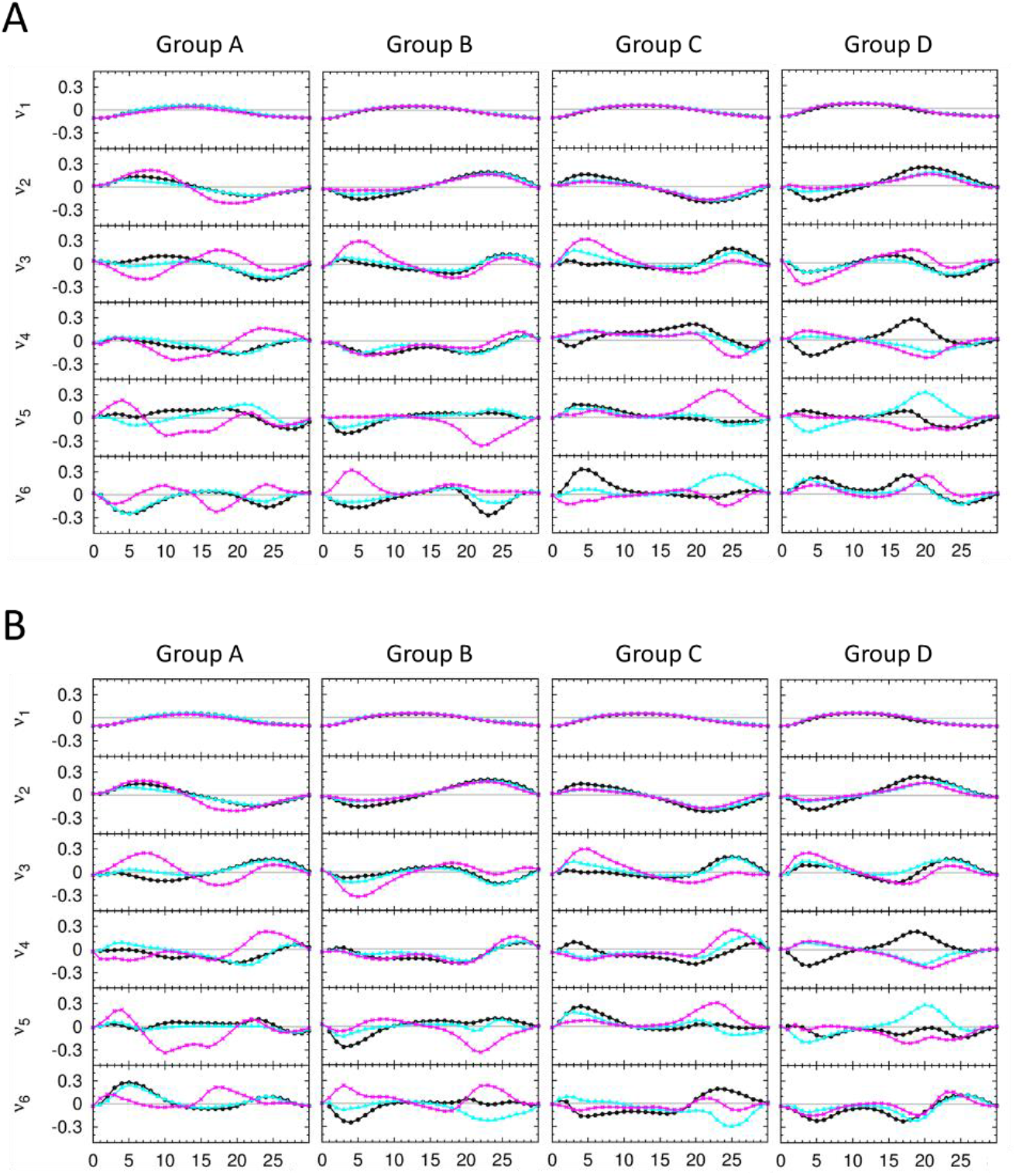
The character of score matrix for each group of average cortical thickness. (A) The results of singular value decomposition analysis on score matrices, which are composed of 365 CN, 481 MCI, 165 AD human brain images as a training set and used for the second iteration of stratified 3-fold cross validation test. And (C) the results of that which are composed of 366 CN, 483 MCI, 166 AD human brain images as a training set and used for the third iteration of stratified 3-fold cross validation test. For each group of average cortical thickness, six singular vectors corresponding to the six largest singular values are presented. Here, x-axis is *m* value defined in the section 2.3, and y-axis is an arbitrary unit for the singular vectors. For each graph, the singular vector components for CN, MCI, and AD subjects are plotted by black, cyan, magenta colors, respectively.

**Supplementary Figure 4.**
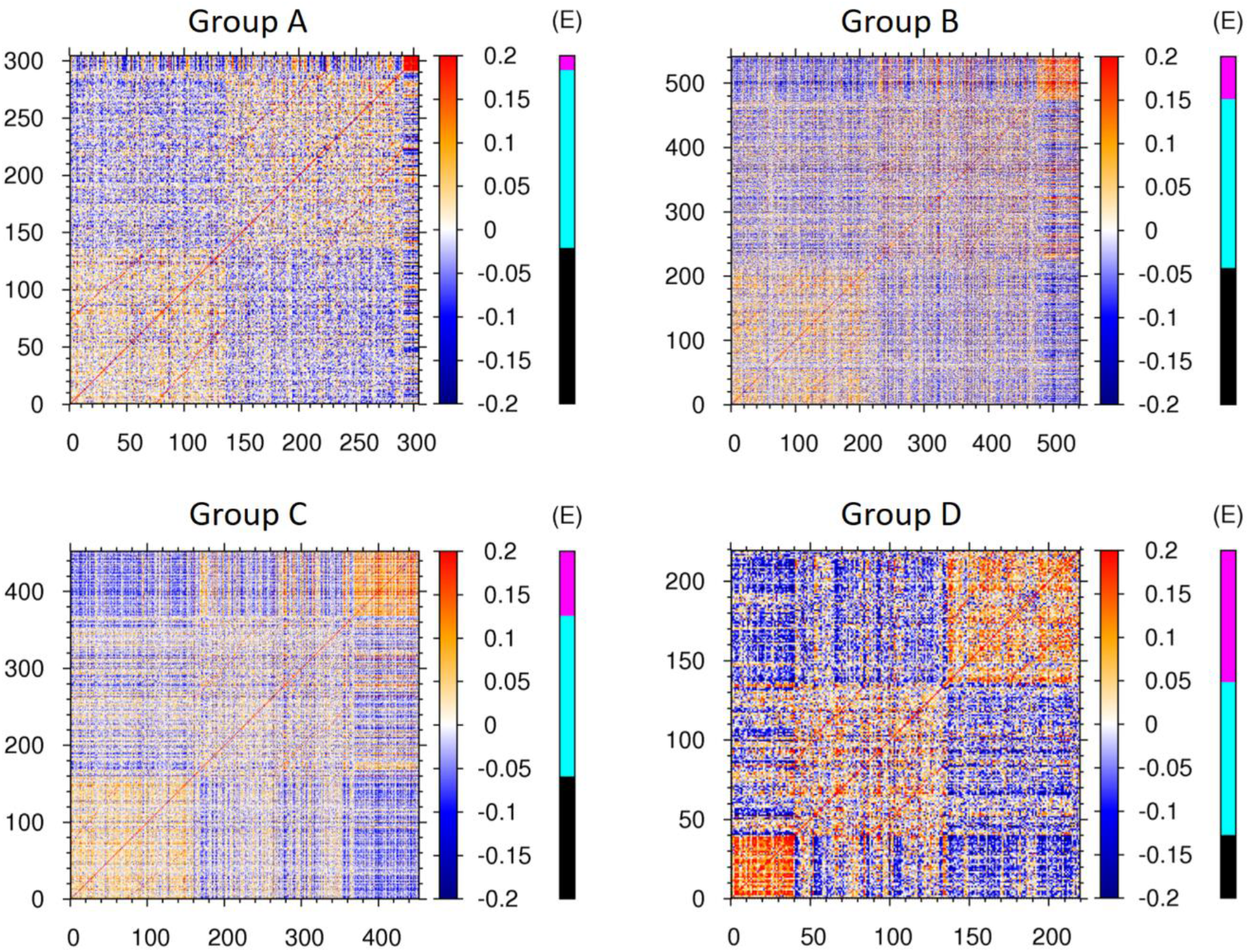
Covariance correlation matrices for each group of cortical thickness. The heat maps is the covariance correlation matrix for group A to D of average cortical thickness. The extra E-cohort color bar at the right of the heat maps represent the clinically determined cohort of cognitively normal subjects and subjects with mild cognitive impairment and Alzheimer’s disease denoted by black, cyan, and magenta colors, respectively.

## References

Dale, A. M., Fischl, B. & Sereno, M. I. 1999. Cortical surface-based analysis. I. Segmentation and surface reconstruction. Neuroimage, 9, 179–94.

Fischl, B., Sereno, M. I. & Dale, A. M. 1999. Cortical surface-based analysis. II: Inflation, flattening, and a surface-based coordinate system. Neuroimage, 9, 195–207.

Galvin, J. E., Sadowsky, C. H. & Nincds, A. 2012. Practical guidelines for the recognition and diagnosis of dementia. J Am Board Fam Med, 25, 367–82.

Hartikainen, P., Rasanen, J., Julkunen, V., Niskanen, E., Hallikainen, M., Kivipelto, M., Vanninen, R., Remes, A. M. & Soininen, H. 2012. Cortical thickness in frontotemporal dementia, mild cognitive impairment, and Alzheimer’s disease. J Alzheimers Dis, 30, 857–74.

Hojjati, S. H., Ebrahimzadeh, A. & Babajani-Feremi, A. 2019. Identification of the Early Stage of Alzheimer’s Disease Using Structural MRI and Resting-State fMRI. Front Neurol, 10, 904.

Im, K., Lee, J. M., Seo, S. W., Yoon, U., Kim, S. T., Kim, Y. H., Kim, S. I. & Na, D. L. 2008. Variations in cortical thickness with dementia severity in Alzheimer’s disease. Neurosci Lett, 436, 227–31.

Kim, B. H., Choi, Y. H., Yang, J. J., Kim, S., Nho, K., Lee, J. M. & Alzheimer’S Disease Neuroimaging, I. 2020. Identification of Novel Genes Associated with Cortical Thickness in Alzheimer’s Disease: Systems Biology Approach to Neuroimaging Endophenotype. J Alzheimers Dis.

Kim, H. J., Ye, B. S., Yoon, C. W., Noh, Y., Kim, G. H., Cho, H., Jeon, S., Lee, J. M., Kim, J. H., Seong, J. K., Kim, C. H., Choe, Y. S., Lee, K. H., Kim, S. T., Kim, J. S., Park, S. E., Kim, J. H., Chin, J., Cho, J., Kim, C., Lee, J. H., Weiner, M. W., Na, D. L. & Seo, S. W. 2014. Cortical thickness and hippocampal shape in pure vascular mild cognitive impairment and dementia of subcortical type. Eur J Neurol, 21, 744–51.

Lebedev, A. V., Westman, E., Beyer, M. K., Kramberger, M. G., Aguilar, C., Pirtosek, Z. & Aarsland, D. 2013. Multivariate classification of patients with Alzheimer’s and dementia with Lewy bodies using high-dimensional cortical thickness measurements: an MRI surface-based morphometric study. J Neurol, 260, 1104–15.

Paternico, D., Manes, M., Premi, E., Cosseddu, M., Gazzina, S., Alberici, A., Archetti, S., Bonomi, E., Cotelli, M. S., Cotelli, M., Turla, M., Micheli, A., Gasparotti R. Padovani, & Borroni, B. 2016. Frontotemporal dementia and language networks: cortical thickness reduction is driven by dyslexia susceptibility genes. Sci Rep, 6, 30848.

Qiu, S., Joshi, P. S., Miller, M. I., Xue, C., Zhou, X., Karjadi, C., Chang, G. H., Joshi, A. S., Dwyer Zhu, S., Kaku, M., Zhou, Y., Alderazi, Y. J., Swaminathan, A., Kedar, S., Saint-Hilaire, M. H., Auerbach, S. H., Yuan, J., Sartor, E. A., Au, R. & Kolachalama, V. B. 2020. Development and validation of an interpretable deep learning framework for Alzheimer’s disease classification. Brain.

Querbes, O., Aubry, F., Pariente, J., Lotterie, J. A., Demonet, J. F., Duret, V., Puel, M., Berry, I., Fort, J. C., Celsis, P. & Alzheimer’S Disease Neuroimaging, I. 2009. Early diagnosis of Alzheimer’s disease using cortical thickness: impact of cognitive reserve. Brain, 132, 2036–47.

Raj, A., Kuceyeski, A. & Weiner, M. 2012. A network diffusion model of disease progression in dementia. Neuron, 73, 1204–15.

Solomon, A. & Soininen, H. 2015. Dementia: Risk prediction models in dementia prevention. Nat Rev Neurol, 11, 375–7.

Tetreault, A. M., Phan, T., Orlando, D., Lyu, I., Kang, H., Landman, B., Darby, R. R. & Alzheimer’S Disease Neuroimaging, I. 2020. Network localization of clinical, cognitive, and neuropsychiatric symptoms in Alzheimer’s disease. Brain, 143, 1249–1260.

Yu, W., Lee, W., Lee, W., Kim, S. & Chang, I. 2011. Uncovering symmetry-breaking vector and reliability order for assigning secondary structures of proteins from atomic NMR chemical shifts in amino acids. J Biomol NMR, 51, 411–24.

